# Precision stratification of risk for suicidal behavior in people with bipolar depression

**DOI:** 10.64898/2026.02.23.26346921

**Authors:** Nina de Lacy, Wai Yin Lam, Mihai Virtosu, Vikrant Deshmukh, Fernando Wilson, Bernice Pescosolido, Ken Smith

## Abstract

Patients with bipolar depression are at the highest risk for suicidal behavior, comprising ∼10% of all deaths. In the critical period preceding attempts, most are not in contact with mental health professionals to effect antisuicidal strategies. There is an urgent need for decision support tools to help nonspecialist providers identify those at elevated risk to facilitate prevention. However, we lack robust, performant predictive models to form the core of such tools. Here, we build a high-precision predictive model of 30-day risk for suicidal behavior using unique electronic health record data from >220,000 patients with bipolar depression. We show that optimized machine learning approaches offer very strong clinical utility, delivering high Standardized Net Benefit in the context of near-perfect calibration and smooth, threshold-robust decision curves. Our results break the longstanding performance ceiling in suicide risk prediction and highlight the importance of training models for clinical utility as well as discriminative skill.

## Introduction

Bipolar disorder is a chronic mental illness characterized by periodic episodes of depression, mania/hypomania and mixed symptoms. Risk for suicidal behavior (SB), or fatal and non-fatal suicide attempts, is higher than in any other psychiatric condition^1^ and 10-20x higher than in the general population,^2^ with 1-year and lifetime prevalence of ∼15% and ∼30% respectively.^3^ Indeed, prior SB for those individuals with bipolar disorder is the strongest predictor of suicide mortality: 15-20% of such patients will eventually die from suicide.^4^ Accordingly, “suicide risk assessment and intervention are invariably at the core of the assessment and management of patients with bipolar disorder.”^5^

Risk for SB in bipolar disorder is state-specific: 70-80% of attempts occur among those with bipolar depression (BD),^6^ where annual prevalence is ∼100x higher than euthymic phases.^7^ However, mitigating risk has proved challenging, evidenced by the stubbornly high prevalence of SB in BD.^7^ A pervasive risk contributor is a lack of treatment engagement amidst system gaps that limit high-risk patients’ access to effective intervention. Among all people with SB, only 1/3 to 1/2 had any contact with mental health services in the year prior to the event.^8^ In bipolar disorder no more than 50% of people who died by suicide were receiving psychiatric treatment and only ∼30% had a mood stabilizer present.^9^ In the critical week prior to the event, only ∼15% visited an Emergency Room or Outpatient Psychiatry.^10^ Thus, the majority of people with BD entering or within periods of near-term SB risk are not in contact with mental health professionals positioned to assess their risk and intervene with anti-suicidal strategies.

In the broader population, screening for suicidal ideation (SI) has therefore been pursued in *nonspecialist* settings such as primary care clinics to improve the chance of detecting people at elevated risk. Clinicians in these settings see ∼45% of those who die from suicide within 30 days prior to the event, double the rate of mental health professionals.^11^ Universal SI screening has been the typical approach but has a number of difficulties. First, it usually captures only current or very recent SI within clinical encounters and is prone to missing individuals who experience risk outside the requisite screening window. Second, all extant screening tools suffer from low discrimination skill.^12^ Finally, limited time and heavy workload are substantial barriers for nonspecialist providers^13,14^ who feel universal screening is cumbersome and can be redundant or produce “overscreening”^15,16^ in the context of perceived low utility. Current recommendations for SI screening are therefore inconclusive^17^ and the need to transition to risk stratification for SB is often highlighted. To do so, more research has been called for into risk stratification tools using electronic health record (EHR) data,^18^ since these are highly scalable and could engage in continuous, encounter-anchored, passive risk monitoring in specialist and non-specialist settings alike to support just-in-time identification without additional burden on clinicians.

Risk stratification is a cornerstone of clinical practice. It mitigates risk by targeting interventions to high-risk patients to improve care, resource allocation and outcomes.^19^ Unlike screening, risk stratification tools are built around predictive models that quantify the degree of individual risk for a specific event over a specific timeframe. SB predictive models with robust clinical utility have not been fully realized due to the inherent difficulties presented by a low base rate and restricted availability of large sample sizes.^17^ An influential 2017 meta-analysis found that prediction skill had not improved over decades and was only slightly better than chance, calling for “a shift in focus … to machine learning (ML) risk algorithms.”^20^ Some ML models have since emerged with better generic discrimination skill (e.g. accuracy or AUROC) than those based in suicide theory.^21^ Yet, their utility is still limited by poor performance in metrics specific to clinical contexts such as Positive Predictive Value (PPV) and Standardized Net Benefit (sNB), even in larger military studies.^22-25^ Such models are less useful in clinical practice since their high false positives needlessly consume provider resources and can result in stigma, bias, alert fatigue and desensitization. Moreover, most SB risk studies do not report clinical utility metrics such as calibration or decision curve analyses.

Constructing effective SB risk prediction models remains an enduring goal. Strategies such as crisis services,^26^ means restriction^27^ and pharmacotherapeutics like lithium,^28^ ketamine^29^ and modern antipsychotics^7^ show evidence for rapid SB risk reduction. A critical gap in advancing effective prevention for SB in BD is constructing a performant, EHR-based risk stratification model. These could be linked with one or more of these interventions to enable continuous risk monitoring and near-term, targeted intervention programs. Such a model does not currently exist, to our knowledge.

We aimed to help close this gap by stratifying individual 30-day risk for SB in patients with BD. We pursued several innovations to construct the first model predicting SB in BD with EHR data. First, we leveraged a historically large sample of >220,000 BD patients. Second, we focused closely on clinical utility and rigor in terms of performance metrics, model optimization and minimization of temporal bias. Thirdly, we compared the relative clinical utility of two complementary approaches: conventional point predictions and longitudinal risk models using our recently-published *RiskPath* toolbox that offers the first explainable AI (XAI) for dynamic prediction.^30^

## Results

### Analytic Strategy

We utilized data from 222,063 patients with a diagnosis of BD in the Epic Cosmos data lake from 2016-2024. Cosmos pools deidentified EHR data from 1,854 participating hospital systems across the United States, representing ∼300 million total patients. We compared point prediction models that used data from a single pre-index encounter and *RiskPath* transformer models that used repeated measures, employing a total of 225 encounter-level and patient-level EHR features (**Supplementary Table S1**). To ensure rigor, all were trained on population-frequency data, tuned via cross-validation and evaluated out-of-sample. Performance was assessed at clinically relevant operating points using PPV at a fixed threshold and within top-k% risk groups, sensitivity, sNB under a policy accepting 10 false positives per true positive, and number needed to evaluate (NNE), alongside global (AUPRC, AUROC) and calibration metrics. Decision-curve analysis (DCA) summarized sNB across thresholds on the same held-out sets. To optimize models, we trialed a cost-sensitive learning variant (penalizing false positives during training), conducted sensitivity analyses across encounter counts (c=2/3/4) and lookback settings (d = 60/120/180 days) and tuned hyperparameters and model structure using principled grid search.

### The 30-day rate of suicidal behavior in patients with bipolar depression was >1% and higher in males

In our sample the 30-day rate of SB was 1.3%, being marginally higher at 1.6% in males versus 1.1% in females.

### Participants with SB had much lower rates of recent participation in mental health care

We analyzed whether participants had attended an outpatient mental health visit up to 3 months prior to the beginning of the 30-day risk window. Among patients without SB, approximately three-quarters had no visits recorded with no appreciable difference between males and females. Among those with SB, <12% had seen a psychiatrist or psychologist prior to their attempt. Rates were slightly lower among males than females (**Table 2**).

**Table 1:** Demographic characteristics of study participants. Demographic statistics of participants with BD included in point prediction models. Patients are allowed to report up to five races in Epic Cosmos and these statistics sum to greater than the total sample size. For statistics in the longitudinal analyses, see **Supplementary Table S2** for the sample sizes and the 30-day SB occurrence rates, and **Supplementary Table S3** for the demographic characteristics of patients.

| Characteristic | Number | Percent |
| --- | --- | --- |
| **All participants** |  |  |
| With SB | 2,803 | 1.3 |
| Without SB | 219,260 | 98.7 |
| **Sex** |  |  |
| Female | 148,751 | 67.0 |
| Number of females with SB | 1,661 | 1.1 |
| Male | 73,185 | 33.0 |
| Number of males with SB | 1,141 | 1.6 |
| Others / did not answer | 127 | 0.1 |
| **Gender** |  |  |
| Woman | 72,939 | 32.8 |
| Man | 32,128 | 14.5 |
| Transgender man | 728 | 0.3 |
| Transgender woman | 583 | 0.3 |
| Others / did not answer | 115,685 | 52.1 |
| **Race** |  |  |
| White | 161,445 | 72.7 |
| Black / African American | 24,151 | 10.9 |
| Asian | 1,562 | 0.7 |
| American Indian / Alaska Native | 881 | 0.4 |
| Native Hawaiian / Other Pacific Islander | 183 | 0.1 |
| Multiple races | 26,669 | 12.0 |
| Others / did not answer | 7,172 | 3.2 |
| **Ethnicity** |  |  |
| Hispanic / Latino | 15,423 | 6.9 |
| Non-Hispanic | 194,802 | 87.7 |
| Did not answer | 11,838 | 5.3 |
| **Age (in years)** |  |  |
| Range | [10.0, 100.0] |  |
| Mean | 45.1 |  |
| Median | 44.0 |  |

**Table 2:**
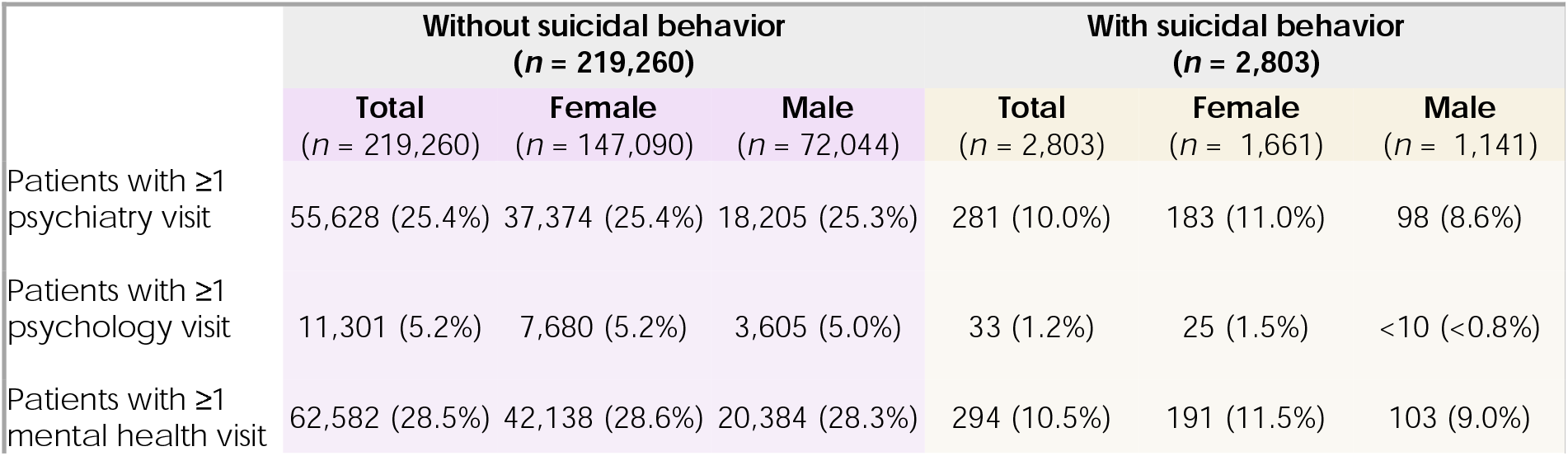
BD Patients with at least one mental health visit within 3 months of risk window. The number of participants in cross-sectional samples for point prediction with at least one psychiatry, psychology or mental health (psychiatry or psychology) visit within three months prior to the beginning of the 30-day risk window. The total counts include patients who did not report their sex. Numbers with less than ten counts are suppressed by Epic Cosmos to protect patient confidentiality. **Supplementary Table S4** shows similar statistics for the longitudinal analyses.

### Point prediction models provided consistently strong and stable high-risk identification across algorithm type

We trialed logistic regression (LR), elastic net (EN: logistic regression with L1/L2 regularization), Support Vector Machine (SVM), gradient-boosted trees via XGBoost (XGB), and artificial neural network (ANN) models. In accordance with contemporary guidelines,^31^ primary performance metrics stressed decision utility for a rare outcome such as SB: PPV, sNB, sensitivity, and number-needed-to-evaluate (NNE) within the top k% risk tranche. These capacity-aligned measures allow clinical managers to pre-specify how many patients they will review, match workload to staffing, keep alert volume stable and minimize alert fatigue. We provide calibration and AUPRC to capture reliability under low prevalence. Complementary secondary metrics support cross-study comparability: sensitivity, balanced accuracy and Youden’s J at 90/95/99% specificity as well as the more generic ML metrics of AUROC and accuracy.

PPV within the top-k% risk groups showed that all training regimes were able to identify enriched high-risk subsets, with close clustering at 0.42-0.51 for 1-2% (**Table 3**). PPV at the global threshold level of 0.5 was quite strong with XGB delivering 0.53 PPV. sNB at top-k% clinical cutoffs reflected the expected monotonic alignment between decision utility and PPV when the number of selected individuals is fixed with strong utility at 0.32-0.60 benefit across all learning regimes. Sensitivity and NNE at k% showed similarly minimal variation. AUPRC, which summarizes ranking performance across the entire score distribution demonstrated similarly minimal dispersion across algorithms as did secondary metrics at high-specificity operating points. Of note, XGB consistently showed marginally superior performance across most primary and secondary metrics but markedly worse calibration statistics, amplified in cost-sensitive learning. Further results may be accessed in **Supplementary Tables S5-S7**.

**Table 3:** Performance of point prediction of SB in participants with BD using EHR data. Performance metrics are shown for models predicting 30-day risk for SB using a single set of EHR features. Features were zero-imputed with additional binary indication for data missingness. The best performance achieved from standard learning is reported. Best models are highlighted in red. All metrics are reported from out-of-sample testing using models trained in data with the actual 30-day SB occurrence rate of 1.3%. ANN = Artificial Neural Network; EN = Elastic Net; LG = Logistic Regression; SVM = Support Vector Machine; XGB = Extreme Gradient Boosting. PPV=Positive Predictive Value; sNB = Standardized Net Benefit; NNE=Number Needed to Evaluate; Bal. accuracy = Balanced accuracy; ECE = Expected Calibration Error; AUPRC = Area Under the Precision Recall Curve; AUROC = Area Under the Receiver Operating Curve. Standardized net benefits were evaluated using a fixed probability threshold corresponding to a 10:1 false-positive-to-true-positive trade-off. Full tables of standard and cost-sensitive learning are shown in **Supplementary Tables S5**.Results are shown after model features are ablated to the point of diminishing performance returns. Results from models obtained with all 211 features prior to ablation may be viewed in **Supplementary Table S6**. Results from models adopted with different data imputation methods may be viewed in **Supplementary Table S7**. The list of features after ablation may be viewed in **Supplementary Table S8**.

| **Primary Metrics** | **ANN** | **EN** | **LR** | **SVM** | **XGB** |
| --- | --- | --- | --- | --- | --- |
| PPV | 0.48 | 0.51 | 0.48 | 0.00 | 0.53 |
| PPV @ top 1% | 0.48 | 0.50 | 0.49 | 0.46 | 0.51 |
| PPV @ top 2% | 0.42 | 0.43 | 0.43 | 0.42 | 0.43 |
| PPV @ top 5% | 0.18 | 0.19 | 0.19 | 0.17 | 0.20 |
| sNB @ top 1% | 0.34 | 0.36 | 0.35 | 0.32 | 0.37 |
| sNB @ top 2% | 0.58 | 0.59 | 0.59 | 0.58 | 0.60 |
| sNB @ top 5% | 0.37 | 0.45 | 0.42 | 0.35 | 0.47 |
| Sensitivity @ top 1% | 0.38 | 0.40 | 0.39 | 0.37 | 0.40 |
| Sensitivity @ top 2% | 0.67 | 0.68 | 0.68 | 0.67 | 0.69 |
| Sensitivity @ top 5% | 0.70 | 0.77 | 0.75 | 0.68 | 0.78 |
| NNE @ top 1% | 2.07 | 1.99 | 2.04 | 2.16 | 1.96 |
| NNE @ top 2% | 2.37 | 2.32 | 2.33 | 2.36 | 2.31 |
| NNE @ top 5% | 5.67 | 5.16 | 5.31 | 5.81 | 5.05 |
| Brier score | 0.01 | 0.01 | 0.01 | 0.01 | 0.01 |
| ECE | 0.00 | 0.00 | 0.00 | 0.00 | 0.01 |
| Calibration Slope | 0.98 | 1.16 | 0.98 | 0.99 | 1.29 |
| Calibration Intercept | 0.00 | 0.04 | 0.10 | 0.04 | -0.32 |
| AUPRC | 0.35 | 0.38 | 0.36 | 0.32 | 0.39 |
| **Secondary Metrics** |  |  |  |  |  |
| Sensitivity @ 99% Spec | 0.67 | 0.67 | 0.67 | 0.67 | 0.67 |
| Sensitivity @ 95% Spec | 0.67 | 0.79 | 0.76 | 0.68 | 0.79 |
| Sensitivity @ 90% Spec | 0.72 | 0.85 | 0.82 | 0.71 | 0.85 |
| Bal. accuracy @ 99% Spec | 0.83 | 0.83 | 0.83 | 0.83 | 0.83 |
| Bal. accuracy @ 95% Spec | 0.83 | 0.87 | 0.85 | 0.82 | 0.87 |
| Bal. accuracy @ 90% Spec | 0.83 | 0.87 | 0.86 | 0.81 | 0.87 |
| Youden index @ 99% Spec | 0.66 | 0.66 | 0.66 | 0.66 | 0.66 |
| Youden index @ 95% Spec | 0.66 | 0.74 | 0.71 | 0.63 | 0.74 |
| Youden index @ 90% Spec | 0.66 | 0.75 | 0.72 | 0.61 | 0.75 |
| AUROC | 0.89 | 0.94 | 0.93 | 0.85 | 0.94 |
| Accuracy | 0.99 | 0.99 | 0.99 | 0.99 | 0.99 |

### Longitudinal prediction achieved reliable discrimination and enrichment, with very high fixed-threshold positive class discrimination

Recent EHR prediction work has increasingly incorporated dynamic models that use repeated measures, including exploratory work in SB.^32,33^ The transformer architecture used here is ideal, because EHR data are sparse and irregularly collected over time.^34^ Across encounter and lookback settings, PPV was consistent within fixed top-k% tranches, indicating reliable enrichment of high-risk individuals at capacity-aligned operating points. sNB at 1% and 2% selections was consistently positive, while at 5% it attenuated and was negative in several configurations, indicating that decision utility concentrated at lower screening fractions (**Table 4**). Sensitivity rose predictably as the selection fraction widened, and NNE mirrored PPV (lowest at 1%, higher at 2% and 5%), supporting operational planning for limited-capacity workflows. PPV at a fixed 0.5 probability threshold was very strong across nearly all configurations at 0.50-0.73 and higher than point prediction. Global and secondary metrics reinforced these patterns. Discrimination was strong across all configurations (AUPRC and AUROC consistently high), with high-specificity operating points (90%, 95%, 99%) yielded uniformly strong sensitivities, balanced accuracies, and Youden indices.

**Table 4:** Performance of longitudinal SB prediction in participants with BD using EHR data. Performance metrics are shown for models predicting 30-day risk for SB using longitudinal EHR features with different numbers of feature encounters (*c* = 2, 3, 4) and width of look-back windows (*d* = 60, 120, 180). Features were zero-imputed with additional binary indication for data missingness. The best performance achieved from standard learning is reported. PPV = Positive Predictive Value; sNB = Standardized Net Benefit; NNE = Number Needed to Evaluate; Cal. Intercept = Calibration Intercept; ECE = Expected Calibration Error; AUPRC = Area Under the Precision Recall Curve; Bal. Acc = Balanced accuracy; AUROC = Area Under the Receiver Operating Curve. Standardized net benefits were evaluated using a fixed probability threshold corresponding to a 10:1 false-positive-to-true-positive trade-off. Full tables of standard and cost-sensitive learning are shown in **Supplementary Tables S9**. All metrics are reported from out-of-sample testing using models trained in data with the actual 30-day SB occurrence rate of 0.4-0.7%. Sample sizes for each model may be inspected in **Supplementary Table S2**. Results are shown after model features are ablated to the point of diminishing performance returns. Results from models obtained with all features prior to ablation may be viewed in **Supplementary Table S10**. Results from models adopted with different data imputation methods may be viewed in **Supplementary Table S11**.The list of features after ablation may be viewed in **Supplementary Table S8**.

| Primary Metrics | c = 2 |  |  | c = 3 |  |  | c = 4 |  |  |
| --- | --- | --- | --- | --- | --- | --- | --- | --- | --- |
|  | d=60 | d=120 | d=180 | d=60 | d=120 | d=180 | d=60 | d=120 | d=180 |
| PPV | 0.50 | 0.59 | 0.51 | 0.62 | 0.73 | 0.60 | 0.68 | 0.65 | 0.63 |
| PPV @ top 1% | 0.38 | 0.41 | 0.41 | 0.34 | 0.32 | 0.35 | 0.26 | 0.25 | 0.29 |
| PPV @ top 2% | 0.22 | 0.24 | 0.23 | 0.18 | 0.17 | 0.19 | 0.15 | 0.13 | 0.16 |
| PPV @ top 5% | 0.10 | 0.11 | 0.11 | 0.08 | 0.08 | 0.09 | 0.06 | 0.06 | 0.07 |
| sNB @ top 1% | 0.47 | 0.50 | 0.50 | 0.55 | 0.51 | 0.55 | 0.44 | 0.41 | 0.52 |
| sNB @ top 2% | 0.41 | 0.46 | 0.45 | 0.42 | 0.34 | 0.43 | 0.28 | 0.23 | 0.36 |
| sNB @ top 5% | 0.08 | 0.12 | 0.14 | -0.12 | -0.12 | -0.06 | -0.34 | -0.43 | -0.24 |
| Sensitivity @ 1% | 0.56 | 0.58 | 0.58 | 0.69 | 0.64 | 0.68 | 0.61 | 0.59 | 0.69 |
| Sensitivity @ 2% | 0.65 | 0.68 | 0.67 | 0.75 | 0.68 | 0.74 | 0.68 | 0.65 | 0.76 |
| Sensitivity @ 5% | 0.75 | 0.77 | 0.78 | 0.82 | 0.80 | 0.82 | 0.75 | 0.71 | 0.84 |
| NNE @ top 1% | 2.66 | 2.46 | 2.44 | 2.96 | 3.13 | 2.84 | 3.83 | 4.07 | 3.40 |
| NNE @ top 2% | 4.59 | 4.23 | 4.27 | 5.41 | 5.95 | 5.18 | 6.86 | 7.48 | 6.18 |
| NNE @ top 5% | 9.92 | 9.39 | 9.16 | 12.45 | 12.55 | 11.68 | 15.53 | 17.06 | 13.90 |
| Brier score | 0.01 | 0.00 | 0.01 | 0.00 | 0.00 | 0.00 | 0.00 | 0.00 | 0.00 |
| ECE | 0.00 | 0.00 | 0.00 | 0.00 | 0.00 | 0.00 | 0.00 | 0.00 | 0.00 |
| Calibration Slope | 0.80 | 1.02 | 0.83 | 0.92 | 0.97 | 0.93 | 0.79 | 0.70 | 0.91 |
| Cal. Intercept | 0.04 | -0.06 | 0.14 | 0.14 | 0.23 | -0.14 | 0.29 | 0.47 | 0.01 |
| AUPRC | 0.34 | 0.38 | 0.35 | 0.50 | 0.46 | 0.47 | 0.39 | 0.38 | 0.46 |
| Secondary Metrics |  |  |  |  |  |  |  |  |  |
| Sensitivity @ 99% | 0.61 | 0.65 | 0.62 | 0.71 | 0.65 | 0.71 | 0.62 | 0.61 | 0.71 |
| Sensitivity @ 95% | 0.75 | 0.77 | 0.78 | 0.85 | 0.81 | 0.83 | 0.76 | 0.72 | 0.84 |
| Sensitivity @ 90% | 0.84 | 0.85 | 0.85 | 0.90 | 0.86 | 0.86 | 0.82 | 0.74 | 0.88 |
| Bal. acc @ 99% | 0.80 | 0.82 | 0.81 | 0.85 | 0.82 | 0.85 | 0.81 | 0.80 | 0.85 |
| Bal. acc @ 95% | 0.85 | 0.86 | 0.87 | 0.90 | 0.88 | 0.89 | 0.86 | 0.83 | 0.89 |
| Bal. acc @ 90% | 0.87 | 0.88 | 0.87 | 0.90 | 0.88 | 0.88 | 0.86 | 0.82 | 0.89 |
| Youden @99% | 0.60 | 0.64 | 0.61 | 0.70 | 0.64 | 0.70 | 0.61 | 0.60 | 0.70 |
| Youden @95% | 0.70 | 0.72 | 0.73 | 0.80 | 0.76 | 0.78 | 0.71 | 0.67 | 0.79 |
| Youden @90% | 0.74 | 0.75 | 0.75 | 0.80 | 0.76 | 0.76 | 0.72 | 0.64 | 0.78 |
| AUROC | 0.93 | 0.94 | 0.93 | 0.95 | 0.93 | 0.94 | 0.92 | 0.88 | 0.95 |
| Accuracy | 0.99 | 0.99 | 0.99 | 1.00 | 1.00 | 1.00 | 1.00 | 1.00 | 1.00 |

### Calibration differentiated usable risk estimates despite similar discrimination performance

Discrimination and calibration address different questions: discrimination asks whether higher-risk patients are ranked above lower-risk patients whereas calibration asks whether the predicted probabilities match observed event rates. In practice, both matter. Good discrimination helps prioritize who should be reviewed first, but safe deployment typically requires well-calibrated absolute risks to align intervention thresholds with real outcomes. In dynamic clinical settings, calibration can drift over time (e.g., shifts in care patterns or coding). Simple recalibration steps are often sufficient to restore alignment. Calibration metrics therefore quantify the reliability of risk estimates and provide a mechanism to maintain reliability under distribution shift.

If we considered discrimination alone, many models would appear similarly strong; calibration clarifies important differences in how usable their probability scales are. In point prediction, our standard learning LR; EN; SVM and ANN models were almost perfectly calibrated with slopes near 1, intercepts near 0, and low ECE (**Table 3**). However, cost-sensitive training often compressed the probability range and shifted intercepts downward, helping explain zero PPV at a 0.5 cutoff in some runs and reduced decision-curve performance (**Supplementary Table S5**). XGB showed a distinct pattern that represented the largest divergence between discrimination and calibration. Under standard learning it was over-confident (slope >1) and this phenomenon is amplified by cost-sensitive learning. Calibration in the longitudinal analyses was similarly generally well behaved: ECE and Brier scores were near zero, slopes clustered close to 1 across configurations, and intercepts showed small, configuration-specific shifts (**Table 4**), with standard learning performing better (**Supplementary Table S5**).

### Decision Curve Analysis

DCA evaluates whether and where a prediction model improves clinical decisions by plotting net benefit across decision thresholds that encode the acceptable trade-off between false positives and true positives. Because DCA is agnostic to prevalence shifts and directly reflects policy choices, it is often the most informative summary of clinical utility. In point prediction, all approaches produced smooth, overlapping curves under standard learning that stayed above the treat-none line across the full threshold range indicating consistent benefit across clinically relevant cut points (**Figure 1**). In contrast, cost-sensitive learning showed early declines and irregular behavior (**Supplementary Figure S1**).

**Figure 1.**
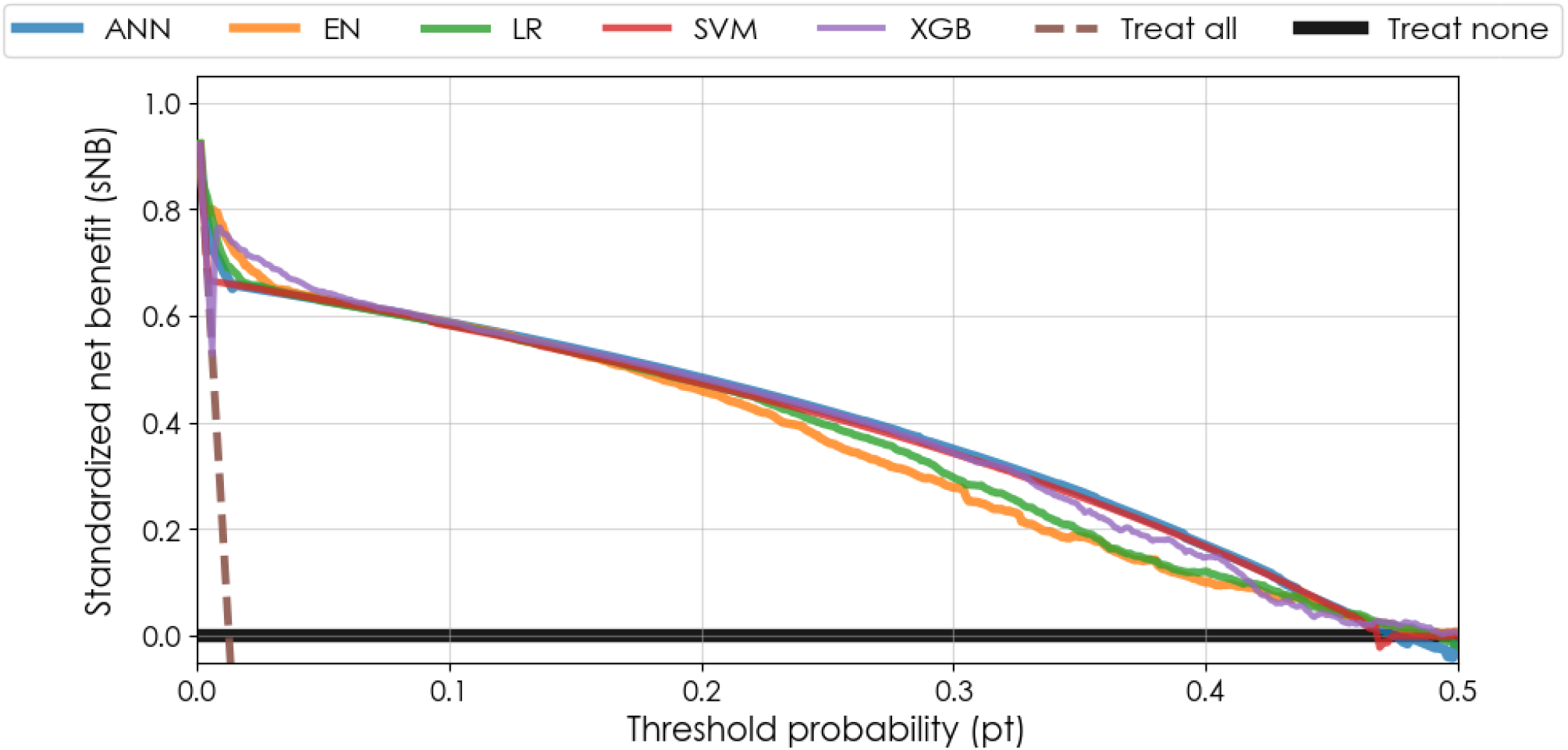
Decision-curve analysis for point prediction models. Standardized net benefit (sNB) is shown as a function of the decision threshold probabilities (p_t_) for five point prediction models (post-ablation) evaluated on the same held-out test set. Features were zero-imputed with additional binary indication for data missingness. The best performance achieved from standard learning is reported. The x-axis is limited to p_t_ ≤ 0.5 to preserve legibility at higher threshold values. The treat-all strategy (brown dashed line) represents the sNB obtained by assigning the intervention to all individuals regardless of predicted risk, while the treat-none strategy (solid black horizontal line) corresponds to withhollding intervention from all individuals. ANN = Artificial Neural Network; EN = Elastic Net; LG = Logistic Regression; SVM = Support Vector Machine; XGB = Extreme Gradient Boosting. See **Supplementary Figure S1** for a similar analysis with the models trained with cost-sensitive learning.

Across lookback windows (60/120/180 days) and encounter requirements (2/3/4), the longitudinal models showed consistent DCA patterns (**Figure 2**). Standard learning produced higher net benefit than cost-sensitive learning across most of the clinically relevant range. Increasing the minimum number of encounters smoothed the very low-threshold region, while changing the lookback length had little effect on the relative ordering of curves. These results are again consistent with calibration results. Taken together, DCA indicates that point-prediction models under standard learning deliver broader and more stable clinical utility across thresholds, while longitudinal models concentrate their benefit at the lowest screening fractions and are more sensitive to training regime and configuration

**Figure 2.**
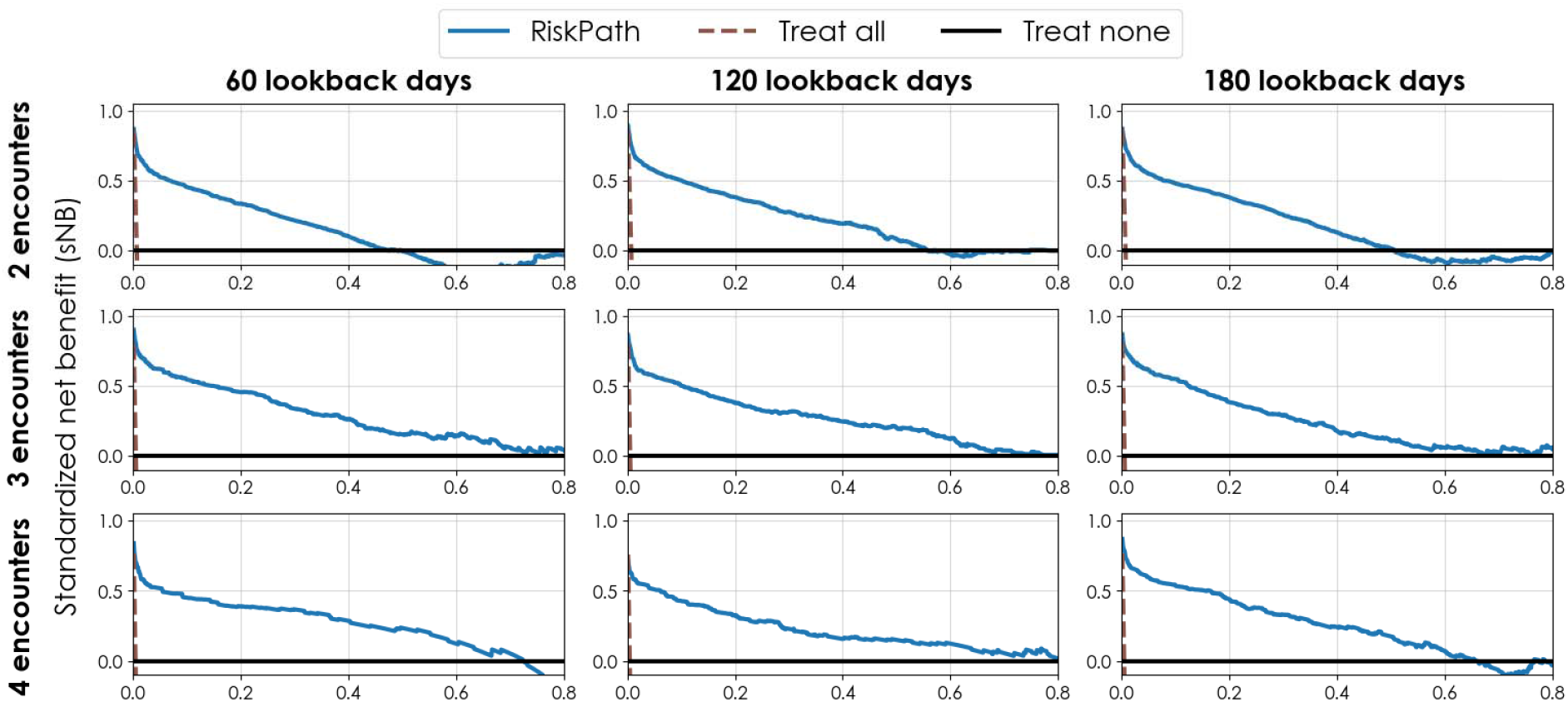
Decision-curve analysis for longitudinal prediction models. Standardized net benefit (sNB) is shown as a function of the decision threshold probabilities (p_t_) for the longitudinal RiskPath models (post-ablation) trained using standard learning (blue). Panels are arranged by the number of encounters (rows: 2, 3, and 4 encounters) and the width of the look-back window (columns: 60, 120, and 180 days). All panels are evaluated on their corresponding held-out test sets. The x-axis is limited to p_t_ ≤ 0.5 to preserve legibility at higher threshold values. The treat-all strategy (brown dashed line) represents the SNB obtained by assigning the intervention to all individuals regardless of predicted risk, while the treat-none strategy (solid black horizontal line) corresponds to withholding intervention from all individuals. See **Supplementary Figure S2** for a similar analysis with the models trained with cost-sensitive learning.

## Discussion

To our knowledge, this study is the first to develop an EHR-based predictive model for SB in patients with BD, an ultra-high-risk population with substantial gaps in continuous mental health care. Our design was carefully formulated to avoid common sources of bias while ensuring that the resultant models could be instantiated in clinical decision tools. In practice, such candidate architectures should next be evaluated in silent mode trials that passively collect real-world data in the EHR and mirror staffing constraints and action costs, with selection based on sNB at the intended operating point and verification of calibration. We approximated the silent trial stage by computing sNB and DCA.^35^ Our results show that all ML architectures were capable of very strong potential clinical utility, delivering 0.30-0.60 sNB in the context of near-perfect calibration and smooth, robust decision curves, with the exception of less ideal calibration in ensemble-based learning. These models can therefore be considered excellent candidates for decision support tools that stratify 30-day risk for SB in BD using point prediction.

SB is a rare outcome, posing substantial modeling challenges due to severe class imbalance and the difficulty of reliably identifying positive cases. To maximize discrimination under these constraints, we evaluated multiple ML algorithms with systematic structural and hyperparameter optimization. Landmarking was integral to this design: by anchoring prediction at outcome-agnostic encounters and restricting predictors to pre-index history, landmarking removed temporal bias. Along with EHR-ready handling of missingness, landmarking renders the model suitable for forward point-of-care use.^36^ An expected trade-off is lower per-encounter prevalence as the at-risk denominator expands. We addressed this by leveraging the nationally representative Epic Cosmos sample of more than 220,000 individuals with BD, which far exceeds sample sizes in prior BD SB prediction studies. The performance achieved here compares very favorably to the historical literature in SB prediction. Prior systematic reviews and meta-analyses show that PPV in earlier ML studies, even in enriched samples, generally ranges from only 6% to 17% despite high values in generic metrics such as AUROC and accuracy.^37^ Against this backdrop, the PPV values obtained in our study substantially exceed these historical ranges, highlighting substantially improved precision and achieving PPV up to 0.53 in point prediction and 0.73 in longitudinal prediction with *RiskPath* using repeated measures. Cost-sensitive learning, often advocated for rare outcomes such as SB, did not meaningfully improve performance and resulted in considerable deterioration to utility, evident in DCA.

Calibration and recalibration are central to safe deployment: calibration may drift with changes in coding, care pathways, or case mix. Simple recalibration is often sufficient to restore alignment, allowing health systems to maintain clinical utility as populations and practices evolve. Despite critiques emphasizing that many existing models are poorly- or uncalibrated or uncalibrated, and calling for calibration as a central evaluation criterion, it has only been performed in <10% of SB predictive studies.^38^ We show that calibration slopes near 1 and intercepts near 0 are natively feasible across most architectures, indicating well-scaled absolute risk estimates suitable for clinical use. Further, calibration helped identify relative weaknesses of the XGB architecture, notwithstanding its marginal advantages in discrimination skill. Taken together, these improvements in PPV, calibration, and decision-analytic net benefit suggest that the models developed here represent a meaningful advance over previous approaches to SB prediction.

Risk-stratification tools can serve different roles along the clinical assessment continuum, and our findings also suggest how to match models and algorithms to use cases. For triage in capacity-constrained settings, models that maximize true positives per unit effort (lower NNE) and provide stable throughput are preferable. Here, the point prediction models met this need by producing a steeper extreme right tail and thus higher PPV at top-k% and fixed specificity, concentrating true cases where actions occur. The longitudinal transformers, by contrast, distributed probability mass across more observations; although they were well calibrated overall and achieved higher unthresholded PPV and strong AUPRC, this dispersion flattened the very top ranks and reduced PPV at 1–5% operating slices. Those same properties, however, can make longitudinal models attractive for earlier, sustained surveillance and low-harm interventions, where cumulative detection over repeated opportunities and overall precision–recall performance matter more than peak top-slice sharpness. Notably, AUROC, accuracy, and balanced accuracy were similarly high across approaches, underscoring that generic metrics are less discriminating for model selection in clinical workflows.

## Conclusions

Our findings show that optimized ML approaches in large data can extract highly meaningful prognostic signals from EHR data to stratify 30-day risk for SB in patients suffering from BD. We identify several model architectures that delivered robust performance comfortably exceeding prior studies in SB to break the longstanding performance ceiling in SB risk prediction. Our results highlight the importance of evaluating models across a diverse set of metrics to ensure that chosen approaches support safe and effective risk SB stratification. In terms of overall clinical and operational utility, our findings lead us to recommend point prediction for its superior performance and easier deployment characteristics given data need only be collected from a single timepoint. Improved calibration metrics, sNB, and PPV-at-k% thresholds support this choice for settings where decisions hinge on well-calibrated absolute risk estimates or low-to-moderate operating thresholds, as is typical for rare event surveillance. More stable calibration slopes and intercepts produced usable probability scales that aligned with realistic risk cut points, translating into consistently higher net benefit across the clinically relevant threshold range. However, longitudinal models may offer advantages in contexts requiring high-specificity identification of a very small number of high-risk individuals, as they yield substantially higher PPV at a fixed generic cutoff, a valuable property in scenarios emphasizing precision over coverage.

### Limitations

Participants may have attended encounters outside the Epic system. EHR and Cosmos data has known biases and limitations, partly due to the data formats supplied from participating health systems. See the *Cosmos Data Domain Encyclopedia* ^39^ for a complete list of known biases and limitations.

## Methods

### Data and Datasets

This work analyzed data from Epic Cosmos, a data repository that consolidates inpatient and outpatient Epic EHRs into a unified patient record. Currently, Cosmos contains records from >300 million patients and 17.6 billion clinical encounters across 1,903 hospitals and 42,400 clinics. Cosmos data undergoes a multi-stage curation and cleaning process managed jointly by Epic and participating health systems. Each site securely transmits data after mapping local values to standardized vocabularies, e.g., International Classification of Diseases (ICD) codes, before transfer to Cosmos. Epic then performs a comprehensive validation process which verifies data parity with source systems, detects mapping errors, and assesses completeness against quality thresholds before approving data for inclusion.^40^ Qualified data are incorporated into the Cosmos relational database, which is refreshed biweekly with updates from all participating sites.

### Study-Specific Data Extraction and Analysis

We extracted the data mart of interest from the Expertly Determined De-Identified database hosted on the Cosmos server, primarily using Structured Query Language queries. The resulting data mart was securely transferred to a project-specific database for analysis. The entire workflow occurred within the Cosmos Data Science Virtual Machine environment, which prohibits exporting line-level or patient-level data to ensure compliance with privacy and security standards. This study was deemed Not Human Subjects reserach by the University of Utah Institutional Review Board.

### Definition of Study Cohort

ICD, Tenth Revision, Clinical Modification (ICD-10-CM) codes were used to classify whether a patient was diagnosed with a specific condition in a given encounter in Cosmos, where an encounter represents different types of interactions with patients (e.g., office visits, inpatient hospitalizations) recorded as a discrete event. Inclusion criteria for the present study were patients who (a) have at least one encounter from January 1, 2016 to December 31, 2024, and (b) had a diagnosis of a current depressive episode of bipolar disorder (ICD-10-CM: F31.3-F31.5) in any of their encounters. These criteria identified an initial cohort of 578,521 patients before further filtering (see below).

### Ascertainment of Suicidal Behavior

The present study defined the clinical outcome, that served as the model’s target label, as the occurrence of SB represented by a binary variable. SB was ascertained using ICD-10-CM codes X71-X83, T14.91, T36-T65, and T71 for intentional self-harm, consistent with the coding framework used in Hansen et al.^41^ Codes indicating subsequent encounters or sequelae (e.g., T14.91XD and T14.91XS) were excluded to avoid misclassifying follow-up care or complications as new SB.

### Temporally Ordered Sampling

To avoid temporal bias, a temporally ordered prediction framework was applied. Temporal bias may occur when information recorded after the outcome is inadvertently incorporated into the definition of the predictor set. This can result in information leakage and inflated prevalence rates and performance estimates that would not generalize well to prospective settings, thereby reducing robustness in deployment. In this study, point prediction models were constructed using information from a single historical encounter per patient, whereas longitudinal prediction models incorporated information from multiple historical encounters. To ensure fair comparison across these settings, a consistent temporally ordered sampling strategy was applied to both model types.

Consistent with established principles for *landmarking* in temporally ordered prediction^5^ a single encounter per patient was designated to determine outcome status, hereafter referred to as the *index encounter*, which defined the timing of the SB outcome. Only encounters occurring strictly before the index encounter were eligible to contribute predictive information and were referred to as *feature encounters*. For controls (patients without SB), an index encounter was analogously designated to serve as a temporal anchor, and all feature encounters were required to occur strictly before this encounter. The number of feature encounters extracted per patient was denoted by *c*. For point prediction models, *c* was fixed at 1, corresponding to the most recent eligible feature encounter prior to the index encounter. For longitudinal prediction models, multiple configurations were evaluated (*c* = 2, 3, and 4) to assess the effect of longitudinal depth on predictive performance. A look-back window *d* was defined as the time interval (in days) between the index encounter and the earliest feature encounter included for a given patient, characterizing how far back potential predictors were traced. Analyses were conducted using *d* = 60, 120, and 180 days.

For subjects in the initial cohort of 578,521, rules were applied to define eligible encounters for outcome forecasting:

(R1) The patient must have at least *c* + 1 encounters, consisting of *c* feature encounters and one outcome (index) encounter.

(R2) The time interval between the index encounter and the most recent feature encounter was greater than 0 days and no more than 30 days.

(R3) For longitudinal prediction models, the time interval between the index encounter and the earliest feature encounter was no greater than the specified look-back window *d*.

(R4) The patient’s age at the index encounter was between 10 and 100 years.

The following procedure was used to operationalize these encounter-level constraints:

- Step 1: Specify the configuration values of *c* and *d*.
- Step 2: For each patient in the initial cohort, designate the patient’s most recent eligible encounter stored in extracted Cosmos data mart as the index encounter, and the preceding *c* encounters as feature encounters.
- Step 3. If Rules (R1) thru (R4) were satisfied, include the patient with the corresponding set of encounters; otherwise, reassign the index encounter to the next most recent encounter of the patient.
- Step 4. Repeat Step 2-3 until the patient was included or had fewer than c + 1 encounters available, in which case the patient was excluded.

The binary target label is identified by the occurrence of SB at the outcome encounter. Sample sizes and prevalence rates resulted from the different configuration of *c* and *d* are presented in **Supplementary Table S1**.

### Features

EHR data typically includes two types of features: patient-level features and encounter-level features. Patient-level features represented characteristics that are fixed or slowly varying over time and not tied to a specific clinical encounter such as demographic characteristics and socioeconomic factors. Encounter-level features captured clinical characteristics that are specific to an individual encounter or to defined look-back periods preceding that encounter such as age, weight, and height at the encounter or healthcare utilization

### Data cleaning and preprocessing

Cosmos explicitly encodes structural missingness (e.g., refusal to answer) and such variables were one-hot encoded as binary indicators representing missingness-not-at-random. Remaining missing values in the extracted dataset were assumed to be missing-at-random. Patient-level variables encoding races were first agglomerated into a single variable then one-hot encoded as the respective race labels. Other nominal variables were one-hot encoded into distinct binary indicators for each value label. Patient-level date variables were encoded as number of days since the patient’s birth date. Encounter-level date variables were encoded as number of days preceding the encounter date. Rare unreasonable negative values (affecting fewer than 21 patients) were identified in certain date-encoded patient-level variables; these were replaced with missing values to reflect unknown data-quality issues. Variables encoding ICD-10-CM codes were first categorized into 22 ICD-10 chapter codes and then one-hot encoded as a list of binary variables, one for each chapter code label. Features with more than 35% missing values were excluded from subsequent analyses. For patient-level variables, missingness was assessed in the standard cross-sectional manner, whereas for encounter-level variables, missingness was evaluated after concatenating all observations across encounters. ML models can perform well with imputation up to 50% missingness, with slightly less lenient thresholds recommended to balance feature retention with imputation risk.

After this filtering process, 124 patient-level features and 101 encounter-level features remained for use. A complete list of variables and their definitions before preprocessing is provided in **Supplementary Table S1**.

Samples were randomly partitioned into training (for modeling fitting) and test (for held-out evaluation) sets using a 70:30 split stratified by the target label. Subsequent preprocessing was performed within each partition to prevent potential data leakage. To remove outliers, continuous variables were clipped at mean ± 3 standard deviations, and ordinal variables were clipped to the valid range defined in the data dictionary. All variables were scaled to [0, 1] using min-max normalization.

To handle missing data, we primarily used zero imputation with an additional binary indicator variable for each feature to explicitly encode missingness. This approach allowed the models to distinguish true zero values and missing observations while preserving potential information contained in missingness patterns. Mean and median imputation strategies were also implemented as sensitivity analyses to assess the robustness of results to alternative imputed values. Prior work in clinical prediction modeling has highlighted the importance of explicitly accounting for missingness patterns using missing indicators.^42^ Results from the mean- and median-imputation sensitivity analyses were reported in **Supplementary Tables S7** and **S11**.

For point prediction the full feature set is simply the union of features from the selected feature encounter, resulting in a single fixed-length input vector. For longitudinal analyses, static patient-level features were broadcast to each encounter and concatenated with the encounter-level features, yielding a three-dimensional data matrix.

### Model fitting

All code used for model development and evaluation is publicly available at: https://github.com/delacylab/Cosmos_Modeling.

ML techniques for point prediction were built with the *scikit-learn* package; XGBoost models were implemented using the *XGBoost* library and ANN models were developed with the *PyTorch* framework.

LR was implemented as a baseline modeling technique, executed with default runtime parameter defined in the *scikit-learn* implementation. EN introduced an additional layer of fine-tuning by performing L1 (LASSO) and L2 (Ridge) regularization to reduce overfitting. *Scikit-learn* provides a built-in tool (*LogisticRegressionCV)* to optimize the regularization strength via cross-validation. SVM models were trained with a linear kernel (*LinearSVC* in scikit-learn) and post-processed with a calibration model (i.e., *CalibratedClassifierCV* in scikit-learn) to generate probability estimates.

XGB models were optimized over number of tree estimators *k* for *k* = 16, 32, …, 512, 1024 via cross-validation to assess how increasing model complexity affected predictive power. ANN models were implemented with a similar cross-validation technique. Each ANN architecture contained three hidden layers, each with *k* hidden units for *k =* 128, 256, …, 896, 1024. Hidden layers are connected by ReLU activation functions to introduce non-linearity with the output layer returning logit estimates. Trainable parameters were initialized using the Xavier uniform distribution.

For longitudinal analysis, we used *RiskPath’s* transformer-based implementation as it enables modeling of sparse temporal information across multiple feature encounters through self-attention while also supporting integrated model explainability via an extension of Shapley Additive exPlanations (SHAP)-based feature attribution for timeseries architectures. Here, each input feature vector was first projected through a linear embedding layer to map the original features into a shared latent space. Positional encodings were then added to the embedded representations to preserve the ordering of encounters in the longitudinal setting. The resulting representations were processed by a multi-layer transformer encoder, which modeled interactions among features across encounters via self-attention. Encoder outputs were aggregated using a global mean-pooling operation to obtain a fixed-length representation for each patient, which was subsequently passed to a fully connected linear layer to produce logit estimates for binary classification. All trainable parameters were initialized using the Xavier uniform distribution. Model width, defined as the dimensionality of the transformer encoder embeddings, was tuned over [128, 256, …, 896, 1024] through 5-fold cross-validation.

For all modeling methods incorporated with a hyperparameter tuning mechanism through cross-validation, the optimal hyperparameter configuration was identified as the one achieving the highest average AUPRC across the cross-validation folds. A final model was retrained with the optimal configuration for held-out evaluation. (**Supplementary Table S12**).

### Evaluation Metrics

We adopted a comprehensive framework for model evaluation based on the STRATOS taxonomy^43^ and following PROBAST guidelines.^31^ This framework allows model performance to be assessed both globally and at clinically meaningful operating points. Model discrimination was evaluated using threshold-independent metrics, including AUROC and AUPRC and threshold-dependent metrics. The latter included PPV sensitivity, specificity, and Youden’s index (sensitivity + specificity – 1). To aid interpretation in low-prevalence settings, we also reported NNE, defined as the reciprocal of PPV, which indicates how many individuals must be screened to identify one true positive.

Beyond the commonly used default decision threshold at 0.5 for binary classification, we evaluated model performance at several clinically relevant operating points. First, specificity-constrained thresholds were defined as the 90^th^, 95^th^, and 99^th^ percentiles of predicted risk among individuals without the outcome. These thresholds were used to define operating points that approximately achieve the targeted specificity levels on the evaluation set. Second, to reflect operational constraints on screening capacity, we evaluated rank-based top-k% operating points (*k* = 1%, 2%, 5%). For each model, individuals in the evaluation set were ranked by predicted risk, and exactly the top *k*% were flagged as positive, ensuring identical flagged samples sizes across models. The rank0based procedure enables fair, capacity-aligned comparisons across models independent of probability scale differences.

Clinical utility was measured net benefit (NB) and sNB. Net benefit was computed as NB = (TP /N) – (FP /N) × [*p*_*t*_ /(1 – *p*_*t*_)] where TP denotes the number of true positives, FP the number of false positives, N the total number of individuals in the evaluation set, and *p*_*t*_ the policy threshold encoding the acceptable FP to TP trade-off. sNB was obtained by normalizing NB by the observevd outcome prevalence. To ensure comparability across models, a single model-independent policy threshold was used when computing SNB (i.e., accepting 10 FPs for each TP).

To evaluate the clinical decision-making utility of prediction models across a range of decision thresholds, we performed DCA following the framework proposed by Vickers and Elkin.^6^ DCA quantifies the NB of using a model to guide intervention decisions by explicitly balancing TPs against FPs as a function of the threshold probability, thereby linking predictive performance to downstream decision consequences. In this work, DCA was applied to models evaluated on the same held-out test sets to enable direct comparison under identical outcome prevalence and decision contexts. We report sNB rather than NB to rescale the y-axis by the outcome prevalence, improving interpretability and comparability of effect sizes across plots.

Calibration was assessed using multiple complementary measures. Slope and intercept were estimated by fitting a logistic regression of the observed outcome on the model’s log-transformed predicted probabilities, with the intercept reflecting systematic under- or over-prediction (ideal value = 0) and the slope reflecting over- or under-fitting (ideal value = 1). Expected calibration error (ECE) used decile-based binning of predicted probabilities, quantifying the average absolute difference between predicted and observed risks across bins. Overall probabilistic accuracy was further evaluated using the Brier score, which measure the mean squared error between predicted probabilities and observed outcomes.

### Cost-sensitive learning

Due to the highly imbalanced outcome distribution, standard unconstrained learning process tends to prioritize the majority class (i.e., controls), thereby increasing the likelihood of FPs. To mitigate the issue, a FP-averse cost-sensitive learning strategy was applied during model training at the loss-function level. Specifically, for each training sample, the standard loss contribution was multiplied by a class-specific weight determined by its label, with negative samples assigned a higher weight and positive samples assigned unit weight. As a result, FP errors contributed more heavily to the total loss and corresponding gradient updates during optimization. This class-weighted loss formulation was applied consistently across all models during training and was not used during evaluation.

By default, the weights for negative and positive samples are both set as 1 to reflect equal weighting between classes in standard learning. To ensure stronger penalties on FPs, a grid search over the negative weights of 2, 3 and 4 was conducted using 5-fold cross-validation. For each candidate weight, models were trained within each training fold and evaluated on the corresponding validation fold, and the optimal weight was selected as the one yielding the highest average recall at the decision threshold corresponding to the 99^th^ percentile of specificity across validation folds. The final model was then refitted on the full training set using the selected weight, together with any other tuned hyperparameters, and evaluated on the held-out test set.

All performance statistics in this paper were reported at models trained with standard learning (i.e., equal class weights). As a sensitivity analysis, performance statistics of the cost-sensitive models were reported in **Supplementary Tables S5-S6** (point models) and **S8-S9** (longitudinal models). Code implementing standard learning and cost-sensitive learning as alternative training configurations was controlled via a runtime parameter in the modeling scripts and was made publicly available in the GitHub repository referenced above.

### Feature ablation

To improve model parsimony and facilitate potential clinical use, a feature ablation procedure was performed for each model. An initial model was trained using the full feature set. *Feature importance scores* were then computed to quantify the relative contribution of individual predictors to the model’s probabilistic output. These scores were used exclusively for feature ranking during ablation.

For LR, EN and SVM, the absolute values of the fitted regression coefficients were used as importance scores to represent feature contribution magnitudes. XGB models natively computed SHAP values and the mean absolute SHAP value across patients was used as the importance score. For ANN and RiskPath transformer models, the *Captum* library^44^ in Python was employed to compute model-specific importance scores via the *GradientSHAP* method, a hybrid of *Integrated Gradients*^*45*^ and *SmoothGrad*,^46^ providing a fast approximation of SHAP values for gradient-based models.

Predictors were ordered by decreasing importance score and models were then retrained and evaluated using the top-*j* predictors for increasing values of *j* with the goal of retaining the most informative predictors while reducing model complexity. The optimal value of *j* was not fixed a priori but was estimated empirically by examining the distribution of feature importance scores. Specifically, a knee-point at the curve was automatically defined identifying the point of diminishing returns in feature importances, defined as the rank beyond which importance scores plateaued. The knee point was identified by the maximum perpendicular length away from the chord connecting the two ends of the importance-rank curve (see **Supplementary Figure S3** for a visualization of this concept).

Feature ablation code including the knee-point technique, based on *kneefinder*^*47*^ is available in our GitHub repository.

## Data Availability

Data used in the present study are available upon application to Epic Systems.

https://cosmos.epic.com/

