## Supplementary figures and images for "Precision stratification of risk for suicidal behavior in people with bipolar depression"

### Supplementary Figure 1

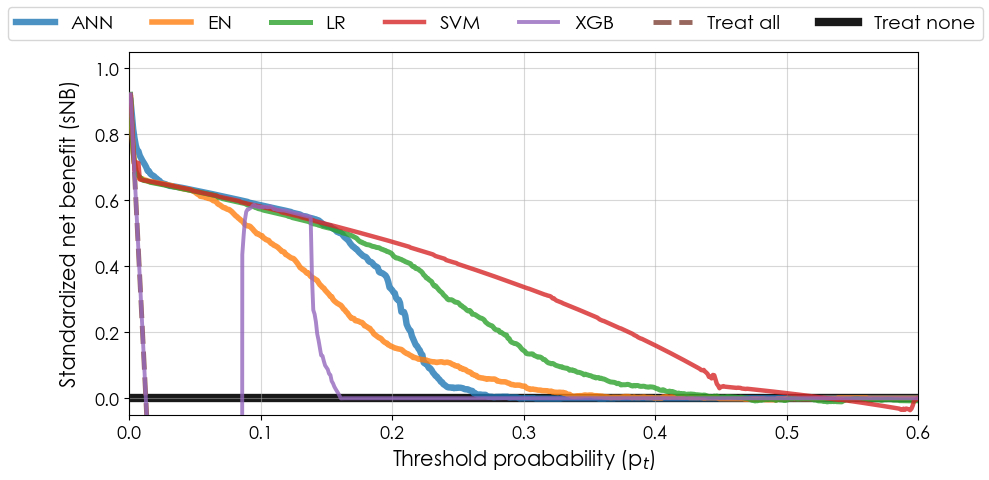

### Supplementary Figure 2

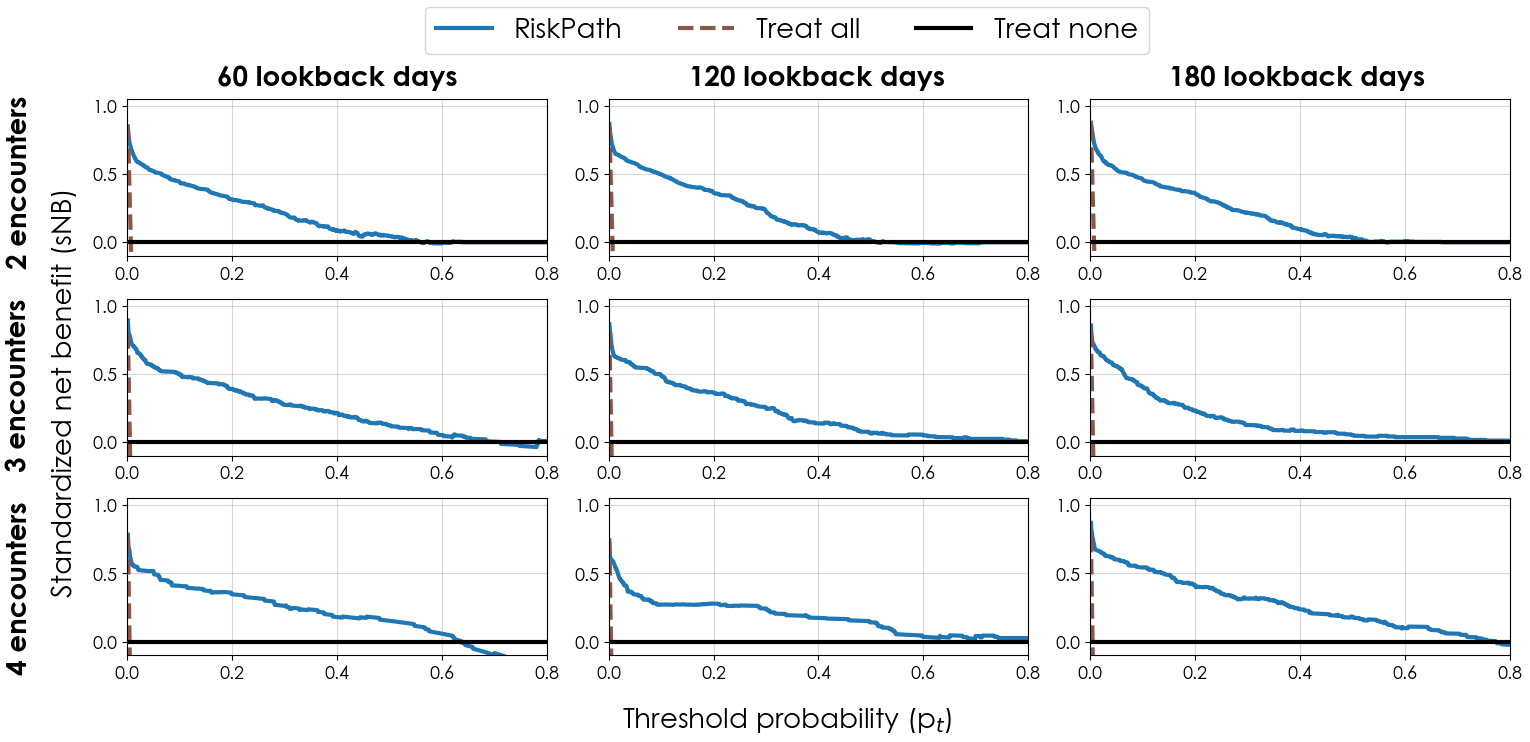

### Supplementary Figure 3

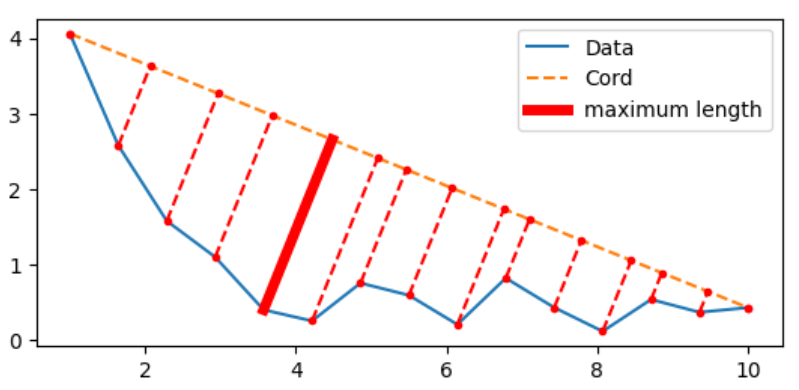
